# BeatAI: BiomEtrics for Atrial Arrhythmia Tracking Using Artificial Intelligence

**DOI:** 10.1101/2025.10.05.25336766

**Authors:** Amin Ramezani, Negin Maddah, Melina Heine, Ali Homaei, Leonard Simeth, Laura Mazuera, Asishana Osho, Jochen D. Muehlschlegel, Jakob Wollborn, Farhad R Nezami

## Abstract

**Background:** Postoperative atrial fibrillation (POAF) affects 20 to 50% of patients undergoing cardiac surgery and is associated with longer hospital stays and adverse outcomes. Although several risk factors for developing POAF have been identified, accurate prediction remains challenging. Wearable ECG patches and remote patient monitoring enable continuous heart rhythm surveillance. Using AI models, subtle yet distinct patterns may be recognized that precede POAF development.

**Objective:** This study evaluates whether combining continuous ECG patch monitoring with deep learning algorithms can improve both early risk stratification and near real-time prediction of POAF.

**Methods:** We analyzed continuous ECG and wearable-derived physiology from 20 postoperative cardiac surgery patients enrolled in a prospective monitoring trial. Each patient wore a 14-day adhesive patch sensor (VivaLNK VV-330) capturing per-second ECG and activity streams. Two complementary deep learning pipelines were developed: (1) a daily-level multimodal Transformer, which downsampled ECG and contextual “TAB tokens” into day-wise units to predict AF occurrence and burden, and (2) an hour-ahead forecasting model, which condensed the last two hours of minute-level physiology into attention-weighted summaries to generate rolling, causal predictions of AF risk in the next hour.

**Results:** Across 162,217 downsampled data elements, the daily-level model showed conservative behavior with very low false negatives, consistently identifying AF-positive days and correctly stratifying high-burden episodes. The hour-ahead forecasting model was trained on 5,607 windows and achieved excellent discrimination (AUC 0.95), high specificity (0.98), and strong predictive value (NPV 0.98). Recall-focused calibration further reduced missed AF hours while maintaining low false alarm rates. Together, the two frameworks provided reliable daily burden stratification and fine-grained, near real-time risk forecasts.

**Conclusion:** Continuous multimodal monitoring combined with AI enables accurate POAF detection, daily risk stratification, and rolling hour-ahead forecasts. This dual-resolution framework has the potential to support perioperative decision-making by enabling earlier intervention, more precise surveillance, and better allocation of preventive therapies in cardiac surgery patients.

**Graphical abstract:** 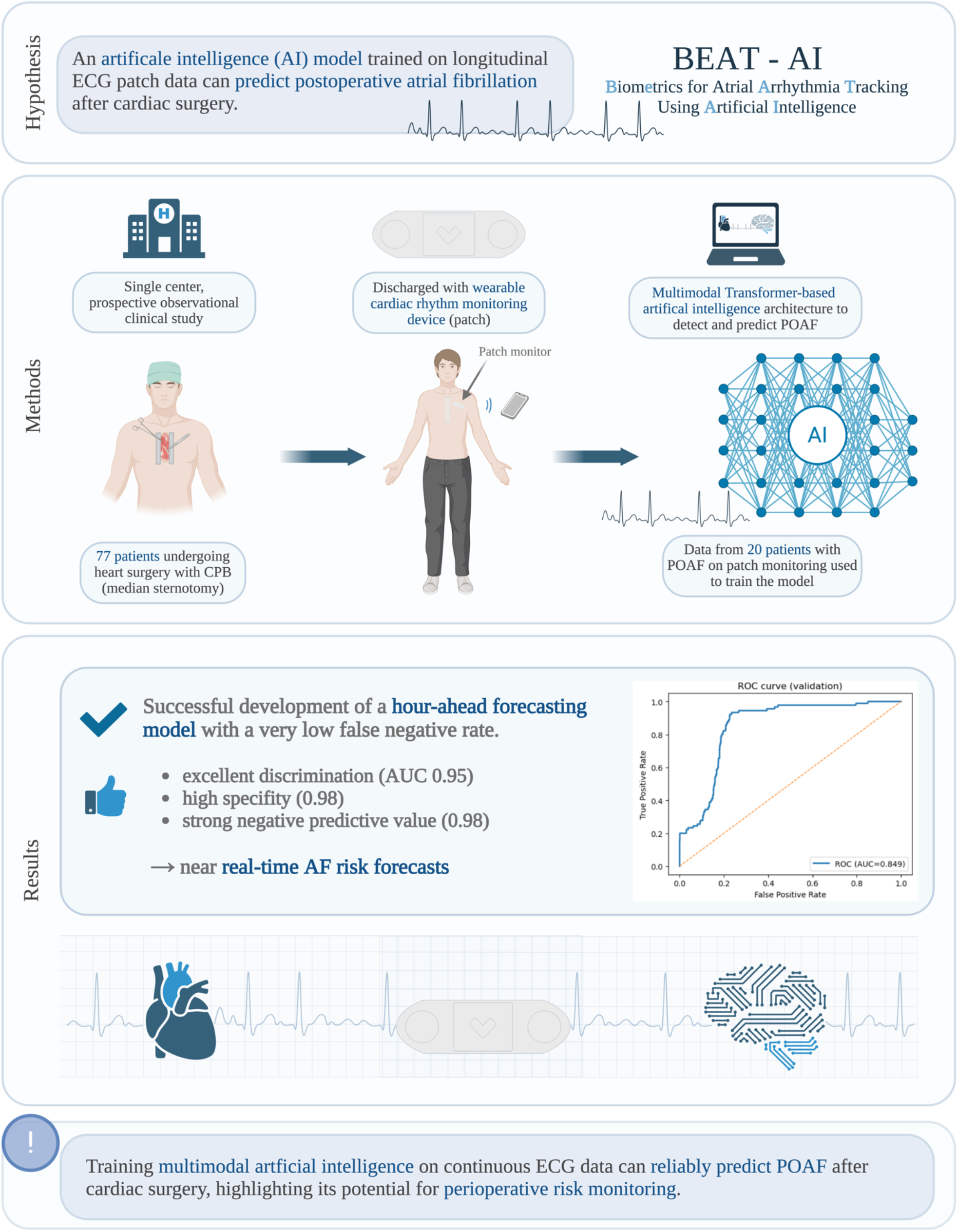

## I. Introduction

Postoperative atrial fibrillation (POAF) is the most common complication after cardiac surgery, with an incidence between 20 and 50% [1], [2]. It is associated with longer ICU and hospital stays, as well as major adverse events, including strokes [3]. Several patient and procedural risk factors have been associated with the development of POAF, including advanced age, chronic kidney disease, heart failure, mitral valve disease and prior atrial fibrillation, as well as surgical factors, such as mitral valve procedures and the use of cardiopulmonary bypass [3], [4]. Despite modern guideline-directed care, accurate risk stratification in the perioperative setting remains challenging: some patients deemed at “high risk” for POAF never experience events, others are not identified early enough for targeted surveillance or preventive therapy [5, 6].

Wearable sensing and ubiquitous ECG acquisition have transformed the healthcare data landscape, demonstrating feasibility for continuous rhythm surveillance and early detection of arrhythmias [7]. Post-discharge remote patient monitoring (RPM) in the cardiac surgical setting has been shown to improve the detection rate of POAF [8]. Continuous cardiac rhythm monitoring technologies generate rich datasets that enable a transition from arrhythmia detection to predictive analysis, by identifying subtle, electrophysiologic signatures that foreshadow incident AF, both outside and within the postoperative context.

Recent advances have shifted from expert-engineered features to deep learning on raw ECG signals. Convolutional and transformer models can extract latent structures, such as premonitory P-wave or atrial substrate changes, and multimodal fusion with imaging and clinical context enhances calibration and generalizability. Self-supervised pretraining further enables strong performance from modest labeled cohorts [9; 10; 11;12]. These methods support earlier AF detection, improved rhythm control strategies, and better stratification for implantable therapies. Yet, key gaps remain, including the need for large-scale validation, prospective trials, and models that are accurate, actionable, and maintainable in real clinical settings [13; 14;15;16].

The present study utilizes continuous ECG data from a prospective remote patient monitoring study and deep learning algorithms to predict postoperative atrial fibrillation (POAF) in cardiac surgery patients. The study aims to evaluate the reliability of using multimodal ECG patch monitoring with deep learning to identify AF episodes and classify arrhythmia burden, thereby enabling earlier and more accurate detection of POAF. We hypothesize that artificial intelligence (AI) models trained on longitudinal electrocardiogram (ECG) patch data can identify subtle electrophysiologic signatures that precede POAF. This, in turn, would result in superior predictive performance compared with conventional risk scores.

## II. Methods

### Study design and setting

We conducted a proof-of-concept analysis on a pilot cohort of postoperative cardiac surgery patients enrolled in an ongoing IRB-approved prospective remote patient monitoring trial at Mass General Brigham (Protocol #2021P000356; NCT04880265). Between January 2021 and October 2024, a total of 100 adult patients undergoing median-sternotomy cardiac surgery with cardiopulmonary bypass and were fitted with an FDA-approved adhesive ECG patch monitor (VV-330, VivaLNK, Campbell, CA) at hospital discharge. In brief, patients were instructed to wear the device continuously for 14 days to capture high-resolution postoperative rhythm and physiologic data. For the present analysis, we focused on a subset of 20 patients, selected as a pilot cohort for developing and evaluating artificial intelligence–based models of postoperative atrial fibrillation (POAF).

### Study cohort and data streams

Eligible patients were adults (>20 years) who provided informed consent. Exclusion criteria included permanent atrial fibrillation (AF), implanted cardiac assist devices, participation in other pharmacological trials and lack of data or poor data quality which could not be analyzed for any heart rhythm in 80% of the postoperative study period. Of the 100 patients enrolled in the parent trial, 77 were included in the primary monitoring analysis (Figure 1). For this secondary AI-focused analysis, we restricted inclusion to patients who experienced at least one documented postoperative AF (POAF) episode lasting ≥30 seconds during the 14-day monitoring period (n=21). One subject was excluded due to poor ECG signal quality, resulting in a final analysis cohort of 20 patients (Figure 1). Continuous physiologic data were acquired from wearable ECG patch sensors placed immediately after hospital discharge. Each record contained a timestamp, short ECG rhythm snippets (128 samples each), and scalar physiologic and activity variables per sample, including heart rate, heart rate variability (RMSSD), skin temperature, activity score, and signal-to-noise ratio (SNR). Accelerometer-based motion summaries were also derived (mean and standard deviation for x, y, and z axes). When available, beat-to-beat intervals (“RRI_array”) and a respiratory surrogate signal (“RWL_array” In addition, pre-sensor clinical variables fixed prior to patch attachment were available for some subjects and incorporated as static covariates. These included age, sex, prior AF diagnosis, history of cardiac surgery, beta-blocker therapy, mitral regurgitation, mitral valve surgery, cardiopulmonary bypass time, and indicators of postoperative ICU or step-down unit care

**Figure 1.**
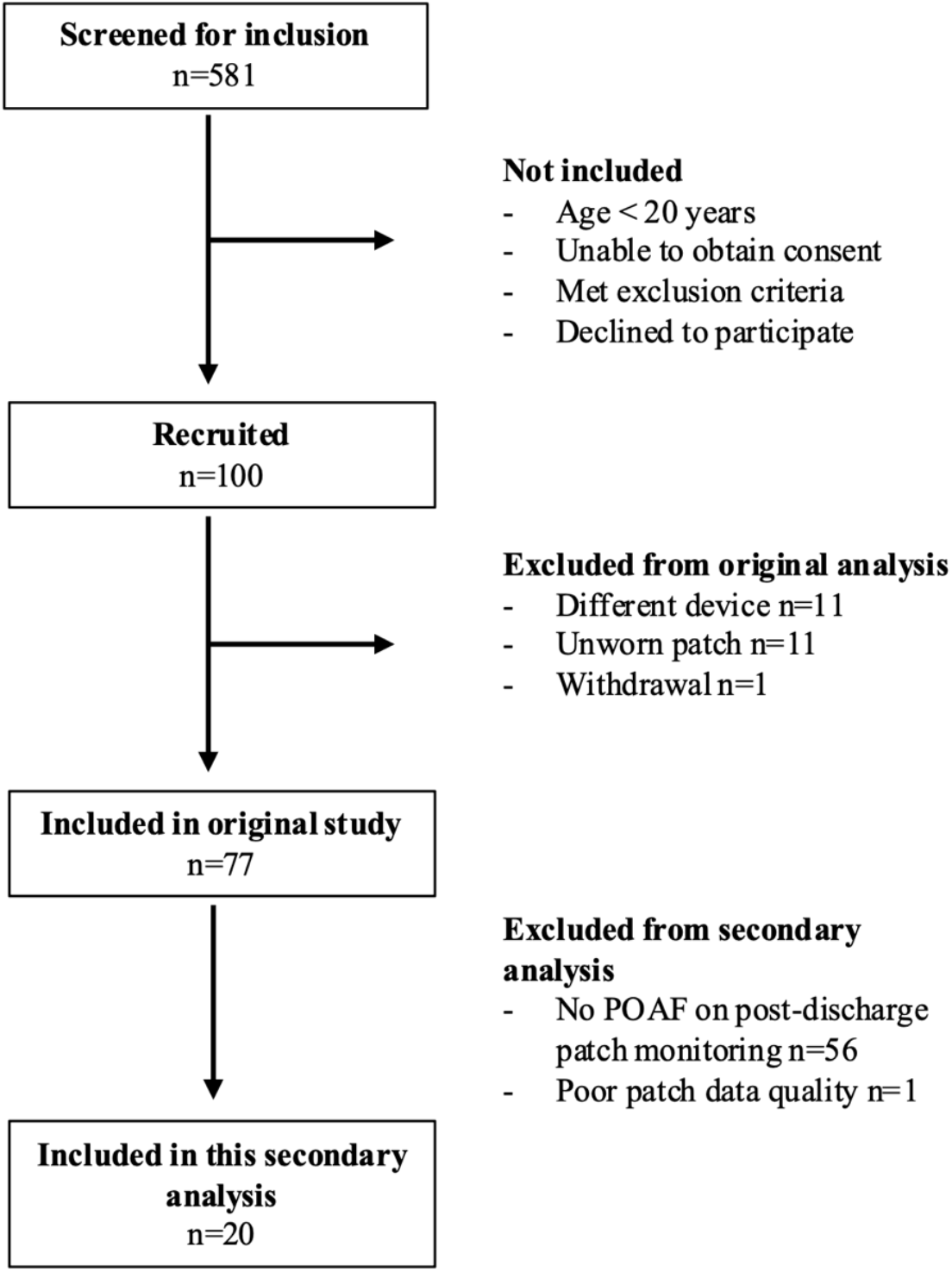
Study flow chart

**Figure 2.**
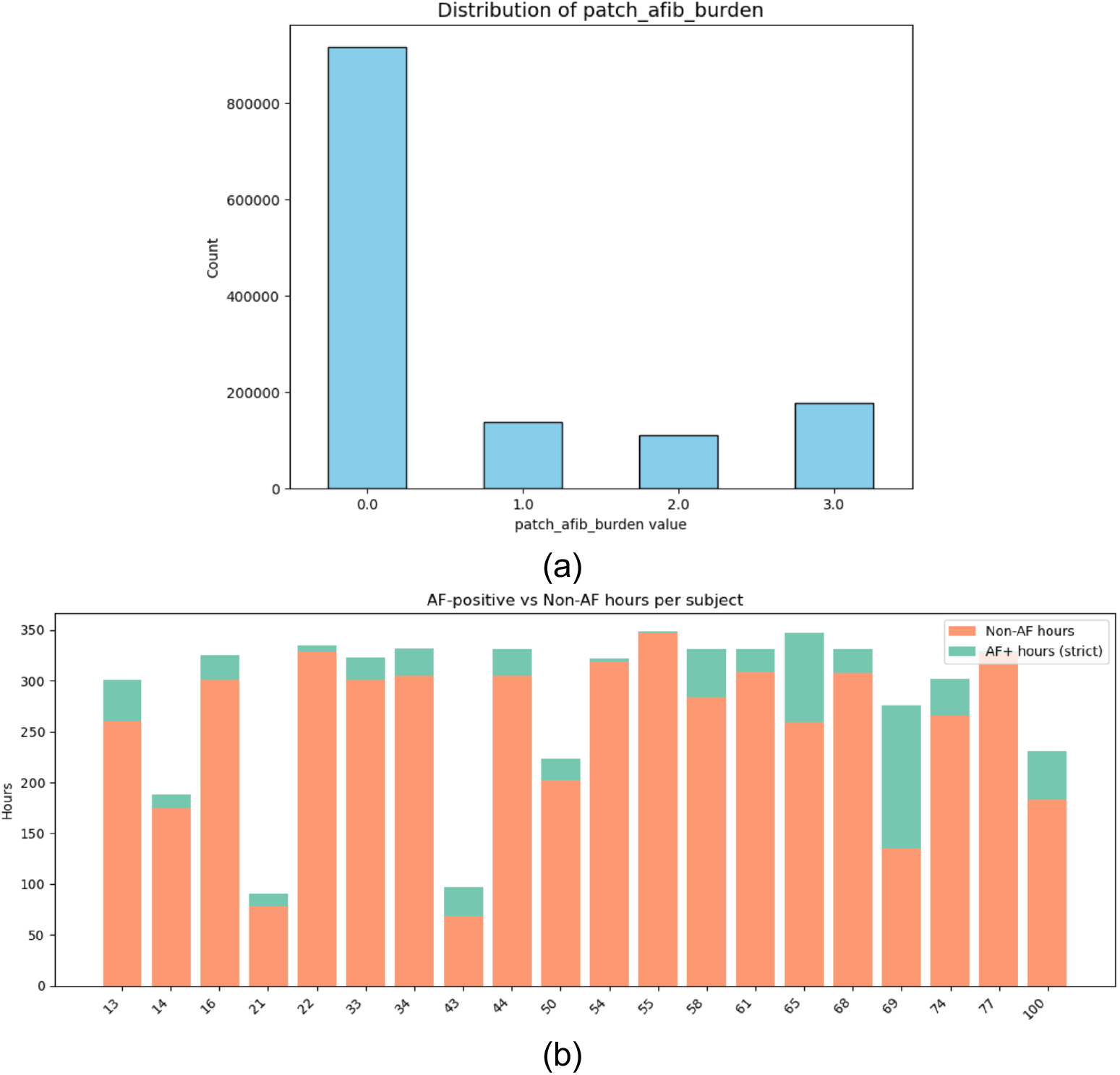
summarizes, per subject, the distribution of AF-positive hours (green) versus non-AF hours (orange), showing the absolute counts of hours labeled AF-positive (strict rule: an hour is AF-positive if it contains ≥5 AF-positive minutes) and non-AF, enabling direct comparison while showing total monitored hours per patient.

**Figure 3.**
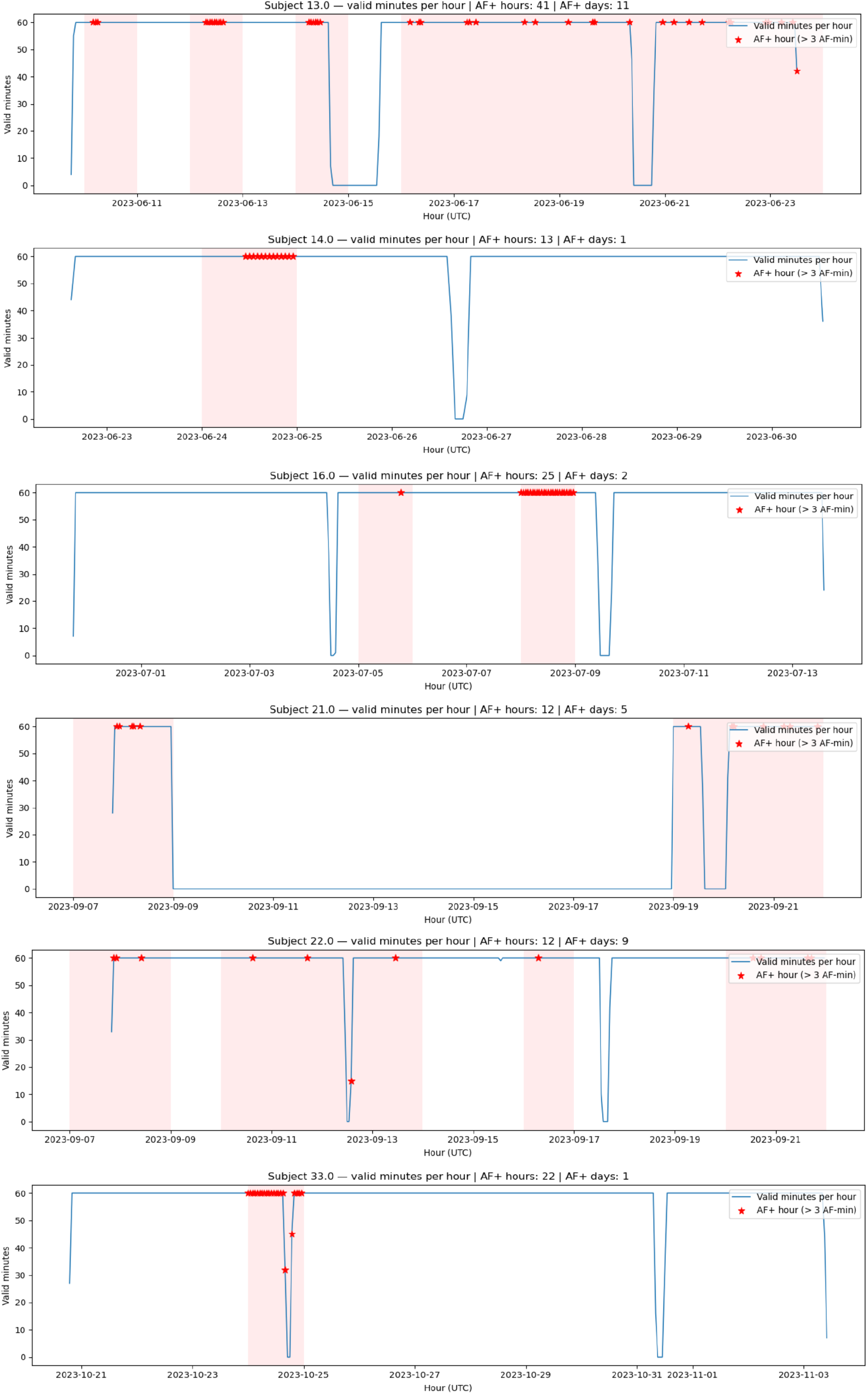
Example time series of valid minutes per hour for two subjects, illustrating atrial fibrillation (AF) burden. Each panel shows hourly counts of valid monitoring minutes (blue line), with AF-positive hours (≥3 AF minutes) highlighted by red stars and shaded regions. Subject 22 (top) experienced 12 AF-positive hours across 9 days, while Subject 33 (bottom) had 22 AF-positive hours concentrated in a single day

**Figure 4.**
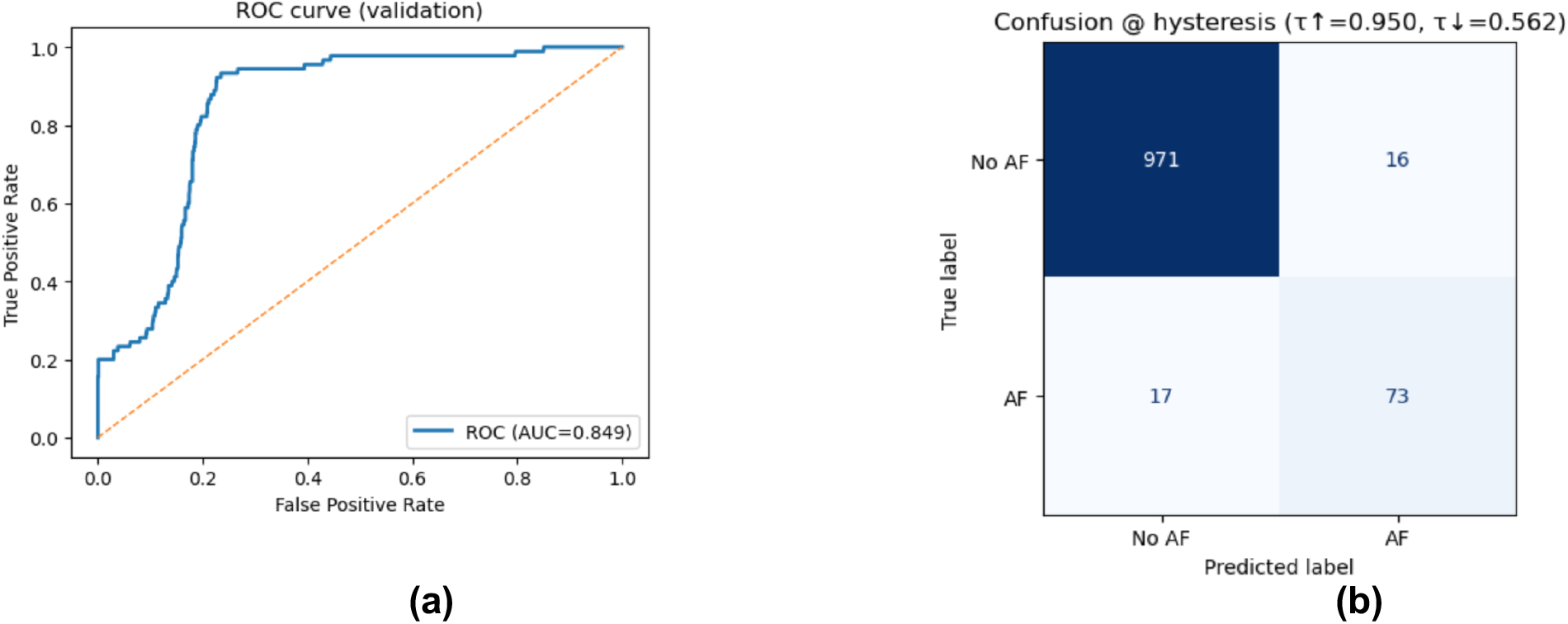
Model performance in atrial fibrillation (AF) detection. *(Left)* Receiver operating characteristic (ROC) curve for the validation dataset, achieving an area under the curve (AUC) of 0.849. *(Right)* Confusion matrix at the optimal hysteresis thresholds (τ↑ = 0.950, τ↓ = 0.562), showing strong discrimination with 971 true negatives, 73 true positives, 16 false positives, and 17 false negatives.

**Figure 5.**
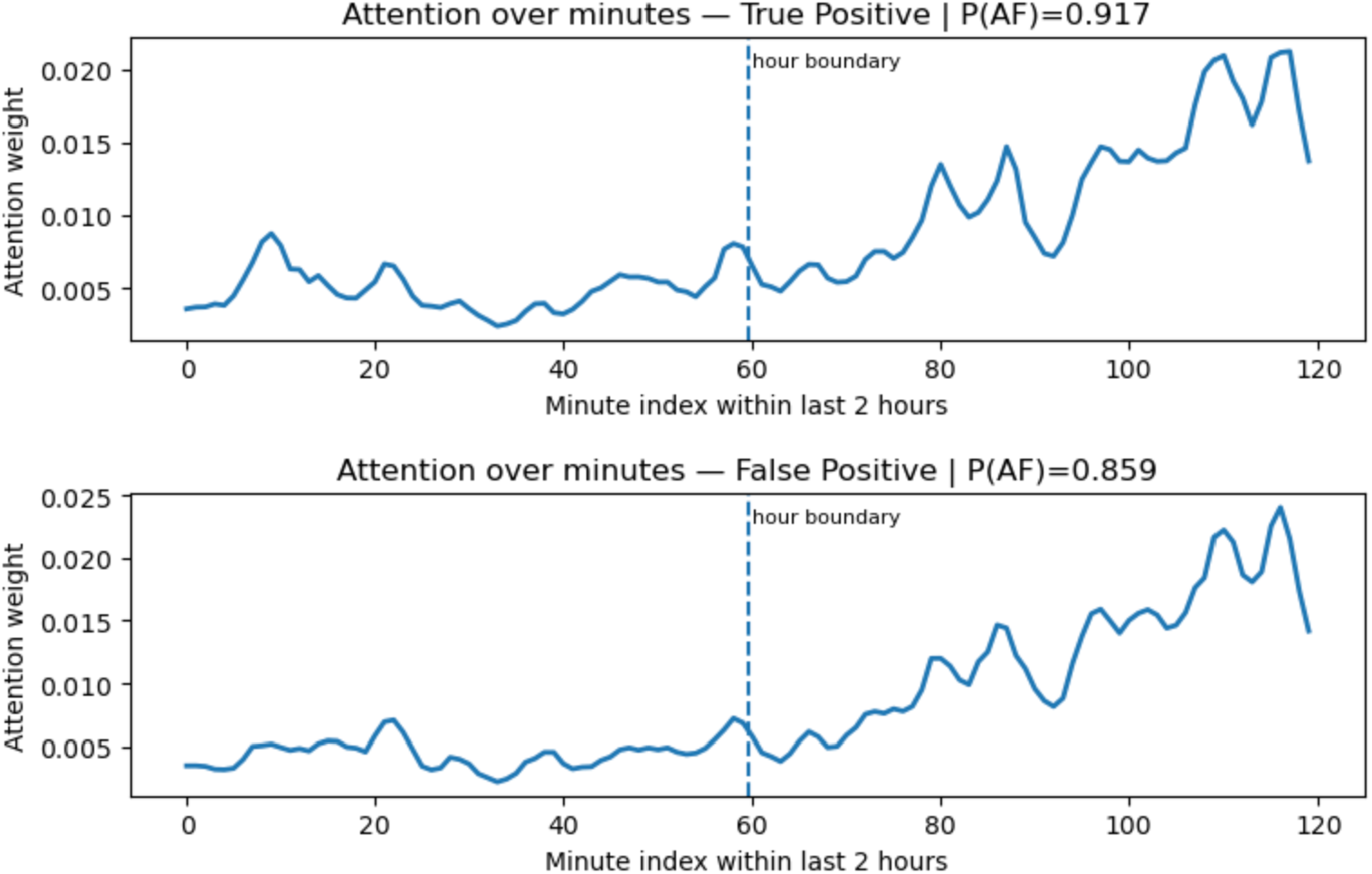
Attention weight distributions over time windows preceding atrial fibrillation (AF) predictions. *(Top)* Example of a true positive prediction (P(AF) = 0.917), where attention increasingly concentrates in the final minutes prior to the AF-positive hour boundary. *(Bottom)* Example of a false positive prediction (P(AF) = 0.859), showing a similar rising attention trend despite absence of AF, highlighting model sensitivity but also potential overfitting to temporal patterns.

### Outcome definitions and prediction task

The study leveraged continuous perioperative monitoring to assess whether advanced artificial intelligence (AI) methods could anticipate postoperative atrial fibrillation (AF) risk across different temporal resolutions. We implemented two complementary prediction frameworks:

#### A) Daily-level risk stratification

Each patient-day was treated as a separate unit of analysis. Continuous ECG and wearable sensor signals were originally recorded at one-second resolution and then downsampled to one sample per minute, yielding 1,440 samples per day. For each minute, a 128-HZ ECG rhythm strip was extracted and paired with its root mean square (RMS) amplitude. In parallel, we engineered tabular context features (“TAB tokens”) that aggregated per-minute physiologic and behavioral measurements, including average heart rate, RMSSD, temperature, motion, activity, respiratory surrogates, and ECG amplitude. Together, these features captured both electrical morphology and physiologic context. A multimodal Transformer model was then trained, projecting ECG and TAB tokens into a shared latent space. The architecture incorporated modality-specific embeddings, day-level positional encodings, masked mean pooling, and cross-modal attention to distinguish true arrhythmia from motion artifacts. The network produced two outputs: (1) binary classification of AF presence versus absence, and (2) ordinal classification of AF burden (single vs. multiple daily episodes). Training followed a two-phase procedure, with the burden head optimized only on AF-positive days. To address class imbalance and computational limits, a sampling strategy retained ∼0.2% of the raw dataset (162,217 rows from 20 patients) while ensuring all AF episodes were represented. Evaluation emphasized clinically relevant metrics: for AF detection, precision, recall, and balanced accuracy, with sensitivity prioritized to minimize missed AF episodes; and for AF burden, recall on high-burden days, given its prognostic significance.

#### B) Hour-ahead forecasting

To enable fine-grained, rolling predictions, we also framed the problem as near real-time forecasting of AF. Hours were labeled AF-positive if they contained ≥5 AF-positive minutes; otherwise, AF-negative. At each time *t*, the model ingested only the preceding two hours of data [t–2h, t) to predict the probability of AF in the following hour [t, t+1h). After an initial two-hour warm-up, the system generated continuous hour-ahead forecasts throughout the 14-day monitoring period, updating in real time as new minutes arrived. For the hour-ahead framework, raw samples were aggregated into minute-level features. For each minute we computed: RMS amplitude of all 128-sample ECG snippets, Vital and activity features (HR, RMSSD, temperature, activity score, SNR), Accelerometer summaries (means and standard deviations of acc_x/y/z), Respiratory surrogates (mean of RWL_array, when available), HRV metrics from RRI_array (mean RR, SDNN, RMSSD, pNN50), A binary AF flag if any sample was AF-positive. Minutes with no valid ECG snippet were discarded. Minutes were then grouped into hours if ≥10 valid minutes were available. For each subject, we constructed sliding 2-hour context windows to forecast the next hour. Additional derived features included: <u>Coverage</u> (observed minutes ÷ 120), <u>Episode history</u> (AF in the most recent hour, run length of the current state, AF count and fraction across the two hours, hours since last AF onset and offset), <u>Circadian terms</u> (sine/cosine of hour-of-day), <u>Relative hour-position scalar</u> (0 for the older hour, 1 for the most recent). When available, static preoperative covariates (e.g., age, sex, prior AF) were appended. To reduce inter-subject variability, minute features were z-scored within each subject’s training set. If insufficient training data existed, a global scaler fit on all training minutes was applied. History, circadian, and hour-position scalars were left unstandardized.

### Model architectures

#### Daily-level multimodal Transformer

The daily model used modality-specific embeddings for ECG and TAB tokens, day-level positional encodings, masked mean pooling, and cross-modal attention. Outputs included binary AF detection and ordinal AF burden classification. Training was staged, with the burden head optimized only on AF-positive days.

#### Hour-ahead hybrid sequence model

The real-time model comprised a three-stage encoder: A) <u>GRU denoising/compression</u> (96 units) to process variable-length minute sequences. B) <u>Transformer encoder</u> (2 layers, 4 heads, GELU activation, width 128) to capture temporal dependencies (e.g., evolving HRV and activity patterns 20–40 minutes earlier). 3) <u>Attention pooling</u> to emphasize informative minutes while down-weighting uninformative or noisy segments.

The pooled summary vector was concatenated with static covariates, coverage, and episode-history features, then passed through two fully connected layers (192→128, ReLU, dropout 0.05). A final linear unit produced a logit, with the sigmoid interpreted as the probability of AF in the next hour.

### Training and validation

Both models addressed class imbalance using weighted binary cross-entropy and focal loss, with oversampling of AF-positive windows. The hour-ahead model used a weighted random sampler to ensure balanced exposure. Optimization was performed with AdamW (learning rate 1×10^−3^, weight decay 5×10^−5^), mixed-precision training, and gradient-norm clipping (1.0). Training was capped at 50 epochs with early stopping triggered by validation plateaus, and learning rates adapted via ReduceLROnPlateau.

To prevent temporal leakage, data were split within each subject: the first 80% of time-series windows were assigned to training and the final 20% to validation. Windows spanning the cutoff were discarded.

### Calibration and decision rules

Because AF episodes often persist once initiated, the hour-ahead system applied a hysteresis decision rule with two thresholds: a start threshold (τ↑) required to switch from non-AF to AF, and a stay threshold (τ↓) required to continue predicting AF once positive. These thresholds were tuned on the validation set through a recall-focused search under a specificity floor (≥0.95, relaxed stepwise to ≥0.85 if unmet). This approach favored minimizing false negatives while keeping false positives low. Model performance was summarized using balanced accuracy, macro-F1, AUC, and confusion matrices.

### Real-time deployment simulation

In simulated bedside deployment, the hour-ahead model continuously generated rolling forecasts after the 2-hour warm-up. Episode-history features were updated based on the model’s own predictions rather than ground truth, mirroring real-world usage. Hours with fewer than 10 valid minutes were automatically flagged as low-confidence. The daily and hourly models thus provided complementary capabilities: robust daily burden stratification and fine-grained, near real-time AF risk forecasting.

## III. Results

We analyzed 20 postoperative patients fitted with continuous patch sensors immediately after surgery. Across all subjects we formed 5,647 hour blocks and constructed 5,607 supervised K-hour windows (K=2 context hours) for learning the next-hour atrial fibrillation (AF) label. After a within-subject temporal split, 4,510 windows were used for training and 1,077 for validation; 20 windows straddling the split boundary were discarded. Using the granular AF signal, AF-positive hours were defined as hours with ≥5 AF-positive minutes. In the validation set, this yielded 90 AF-positive and 987 AF-negative hours (AF prevalence ≈ 8.4%). (Descriptive burden context across the dataset: unique AF burden values were {0,1,2,3} with counts 917,546; 136,868; 109,362; and 177,275 rows respectively; by subject, 18 had any 0-burden days, 14 had 1-burden, 11 had 2-burden, and 7 had 3-burden days.)

At the neutral decision thresholds (τ↑ = τ↓ = 0.5), the model demonstrated strong discriminatory ability on the validation set, achieving an area under the ROC curve (AUC) of 0.947, which reflects excellent discrimination. The balanced accuracy at this point was 0.883, with a macro-F1 score of 0.835. The corresponding confusion matrix was [[TN = 942, FP = 45], [FN = 17, TP = 73]], translating into a sensitivity of 0.811 (73 out of 90 AF-positive hours correctly identified) and a specificity of 0.954 (942 out of 987 AF-negative hours correctly identified). Recognizing that missed AF episodes are clinically more costly than false alarms, we applied a recall-focused hysteresis calibration strategy. This involved tuning two separate thresholds: a higher start threshold (τ↑) to switch from no-AF to AF predictions, and a lower stay threshold (τ↓) to maintain an AF prediction once already positive. Using a recall-constrained search with a specificity floor of 0.95, the optimal operating point was identified at τ↑ = 0.931 and τ↓ = 0.183. At this calibration, the model achieved a confusion matrix of [[TN = 951, FP = 36], [FN = 15, TP = 75]], yielding a sensitivity of 0.833, a specificity of 0.964, a precision of 0.676, and a recall-weighted F_2_ score of 0.796. This adjustment reduced false negatives from 17 to 15 while simultaneously lowering false positives from 45 to 36 compared with the neutral setting. For the final reporting, we adopted a slightly more conservative configuration, fixing the start threshold at τ↑ = 0.931 and increasing the stay threshold to τ↓ = 0.600. This choice reflects a preference for sustaining AF alerts only when there was moderate ongoing evidence, thus reducing the likelihood of unnecessary prolonged alerts. At this final operating point, the model achieved a balanced accuracy of 0.897, a macro-F1 score of 0.899, and an overall accuracy of 0.969. The confusion matrix was [[TN = 971, FP = 16], [FN = 17, TP = 73]], corresponding to a sensitivity of 0.811, a specificity of 0.984, a positive predictive value (PPV) of 0.820, and a negative predictive value (NPV) of 0.983.

The system therefore missed 17 AF hours (18.9% of AF hours) and generated 16 false alarms (1.6% of non-AF hours) in validation. Discrimination remained strong (AUC as measured pre-threshold 0.947). A strong baseline that predicts “next hour equals last hour” achieved balanced accuracy = 0.897 and macro-F1 = 0.897. The calibrated model matched balanced accuracy and slightly improved macro-F1 (0.899), indicating it learned informative patterns beyond rhythm persistence while maintaining very high specificity.

## IV. Practical implication

These results support prospective, rolling hour-ahead AF forecasts after only a 2-hour warm-up, with excellent specificity and high NPV (98.3%). Depending on clinical priorities, the hysteresis thresholds can be shifted toward higher sensitivity (recall-focused pair: τ↑=0.931, τ↓=0.183) to further reduce missed AF at acceptable false-positive rates. The model behaved conservatively, with very low false negatives on AF-negative days. Occasional false positives occurred, but importantly, all high-burden AF days were correctly identified, an acceptable trade-off for clinical monitoring.

## V. Discussion

Remote patient monitoring using wearable ECG technology has been shown to be feasible and effective in enhancing the detection of postoperative atrial fibrillation (POAF) after cardiac surgery [5]. Building on this foundation, our study demonstrates that predictive modeling can extend the value of monitoring by moving beyond retrospective detection toward proactive, real-time risk stratification. Using a multimodal Transformer architecture at the daily level and a hybrid GRU–Transformer model at the hourly level, we showed that artificial intelligence (AI) can capture both long-term burden signatures and short-term temporal dynamics of AF onset. Several important insights emerge from our findings. First, the attention mechanism in the hour-ahead forecasting model consistently emphasized the most recent hour of patient data as the strongest predictor of AF onset. This highlights the clinical relevance of short-term fluctuations in physiology: subtle changes in heart rate variability, ECG morphology, or activity levels immediately preceding an episode carry greater predictive weight than more distant history. By structuring the task at the hour-level granularity, our framework produced rolling forecasts that updated continuously after a brief warm-up, demonstrating how machine learning can transform continuous monitoring into actionable, real-time risk assessments that reflect the evolving physiologic state of the patient.

At the same time, the daily-level multimodal Transformer complemented this approach by reliably distinguishing AF-positive from AF-negative days and stratifying burden (single vs. multiple episodes). The model’s conservative behavior, with very low false negative rates, ensured that AF-negative days were rarely misclassified. From a clinical perspective, this property is desirable: minimizing missed AF episodes is more critical than eliminating every false alarm. Timely recognition of arrhythmia burden enables early interventions, such as antiarrhythmic therapy, anticoagulation, or closer outpatient surveillance, that may reduce risks of stroke, heart failure, or rehospitalization. Importantly, the architecture leveraged cross-modal interactions: irregular ECG morphologies coinciding with motion artifacts were downweighted, while physiologically consistent irregularities were emphasized. This joint modeling of ECG and contextual signals underscores the value of multimodal fusion in perioperative risk monitoring.

From a healthcare systems perspective, the hour-ahead model bridges the gap between passive monitoring and proactive intervention. By predicting AF risk on a rolling basis, the system creates opportunities for clinicians to intervene earlier, rather than relying solely on retrospective burden measures. This capability empowers both physicians and patients with data-driven awareness of evolving risk profiles during recovery. Moreover, the methodological contributions, attention-based recurrent encoders, hysteresis thresholds, and recall-focused calibration, offer a scalable blueprint for forecasting other time-sensitive complications in critical care.

Despite these encouraging findings, several limitations need to be acknowledged. The analysis was restricted to a small cohort of 20 patients, which limits generalizability. While downsampling strategies enabled efficient training, external validation in larger and more diverse populations is essential to confirm robustness. Prediction of single-episode AF days remains challenging, reflecting the intrinsic difficulty of distinguishing brief paroxysms from sustained arrhythmia. Finally, while cross-modal attention and feature weighting provide some interpretability, more formal explainability techniques will be needed to foster clinician trust and support regulatory approval. Taken together, our results demonstrate that integrating continuous multimodal ECG monitoring with advanced AI architectures enables both reliable detection and meaningful prediction of POAF. The dual-resolution framework, daily-level burden stratification and rolling hour-ahead forecasting, illustrates how AI can be embedded into perioperative workflows to support earlier intervention, better risk stratification, and ultimately improved outcomes for cardiac surgery patients.

## VI. Conclusion

This pilot study demonstrates the feasibility of using multimodal artificial intelligence (AI) methods, combining ECG signals with wearable-derived physiologic features, to predict postoperative atrial fibrillation (POAF) in cardiac surgery patients. By implementing both a daily-level multimodal Transformer and an hour-ahead hybrid sequence model, we showed that AI can capture complementary patterns of arrhythmia risk: robust identification of AF presence and burden across patient-days, and fine-grained, rolling forecasts that anticipate AF onset with high specificity in near real time. Despite the small sample size, both models consistently minimized missed AF episodes, a clinically desirable property in perioperative monitoring where the cost of false negatives outweighs that of occasional false alarms. The framework thus aligns with real-world needs, offering both reliable burden stratification and actionable short-horizon risk prediction. Future work will focus on scaling to larger, multi-center cohorts, external validation to ensure generalizability, and advancing interpretability to build clinician trust and regulatory readiness. With these developments, multimodal AI systems have the potential to evolve from passive monitoring tools into proactive decision-support platforms, enabling earlier intervention, more precise risk stratification, and ultimately improved outcomes for patients recovering from cardiac surgery.

## Data Availability

All data produced in the present study are available upon reasonable request to the authors

